# Spinal cord stimulation restores sensation, improves function, and reduces phantom pain after transtibial amputation

**DOI:** 10.1101/2022.09.15.22279956

**Authors:** Ameya C. Nanivadekar, Rohit Bose, Bailey A. Petersen, Elizaveta V. Okorokova, Devapratim Sarma, Juhi Farooqui, Ashley N. Dalrymple, Isaiah Levy, Eric R. Helm, Vincent J. Miele, Michael L. Boninger, Marco Capogrosso, Sliman J. Bensmaia, Douglas J. Weber, Lee E. Fisher

## Abstract

In the United States, over 1.5 million people live with lower-limb amputation. Existing prosthetic limbs do not restore somatosensory feedback from the limb, resulting in functional impairments including balance deficits and an increased risk of falls. Further, these prostheses do not alleviate the severe phantom limb pain that often follows amputation. Leveraging clinically available spinal cord stimulation electrodes, we designed a system that restores somatosensation in the missing limb, thereby improving balance and gait and reducing phantom limb pain. We show that spinal cord stimulation can evoke sensations in the missing foot and that we can control the location and intensity of those sensations. Further, by modulating stimulation intensity in real time based on signals from a wireless pressure-sensitive shoe insole, subjects exhibit improvements in functional measures of balance and gait stability. Finally, over the duration of the implant period, subjects experienced a clinically meaningful decrease in phantom limb pain. These combined results demonstrate that, with an electrode technology that is currently in widespread clinical use, our approach has the potential to become an important intervention for lower-limb amputation.

## MAIN

Every year, approximately 150,000 people in the United States undergo amputation of a lower limb^1^. Loss of a lower limb leads to chronic challenges including major mobility impairments and emergence of chronic pain that appears to emanate from the missing limb (i.e. phantom limb pain, PLP). Current clinical practice involves prescribing a prosthetic limb to improve functional mobility, along with neuroleptic and opiate pharmaceuticals to treat PLP. Even with these interventions, people with lower-limb amputation exhibit a high rate of falls, a lack of confidence during gait, abnormal gait patterns, and persistent PLP. All these problems have been associated with the disruption of somatosensory feedback from the missing limb. Tactile feedback from the sole of the foot is critical for maintaining balance and postural stability^2^ and the loss of somatosensory feedback after an amputation causes a sensorimotor mismatch between attempted movements and expected sensory feedback. This mismatch has been implicated in the development and maintenance of PLP^3^. One potential way to address the sequelae of lower-limb amputation is to restore somatosensation in the missing limb, thereby improving functional outcomes, and reducing PLP.

Previous studies have demonstrated that electrical stimulation of peripheral nerves in the residual limb can evoke sensations in the missing hand or foot^4–6^. Tactile feedback via peripheral nerve stimulation has been shown to enhance control of the prosthesis and improve balance and gait^7–11^. Additionally, anecdotal evidence suggests that chronic peripheral nerve stimulation reduces PLP^7,12–14^. To date, most studies to restore somatosensory feedback from the missing limb have relied on complex surgical techniques to implant devices inside or around peripheral nerves or to reroute those nerves to other regions of the body^6,7,9,11,15,16^. While these approaches clearly demonstrated the promise of electrical stimulation, their surgical complexity remains a barrier to widespread clinical adoption. Evoking sensations via peripheral nerve stimulation may also be challenging in individuals with severe peripheral neuropathy, a common co-morbidity for people with amputations related to vascular disease and diabetes, which account for up to 82% of lower limb amputations^17^. To our knowledge, no study to date has demonstrated restored somatosensation in the amputated foot for people with diabetic amputation.

Here, we aimed to address these challenges by leveraging spinal cord stimulation (SCS), rather than peripheral nerve stimulation, to restore somatosensory feedback from the missing lower limb. SCS is an existing clinical technology that is implanted in as many as 50,000 people each year to treat chronic pain^18^. The surgical procedures involved in the implantation of these devices and the associated risks are well understood, and most major medical centers throughout developed countries have physicians that routinely perform SCS implants^19^. Recently, we have shown that cervical SCS can be used to restore somatosensation from the missing hand in people with upper-limb amputation^20^. Our goal in this study was to demonstrate that lumbar SCS could evoke sensations in the missing foot, and that the restored somatosensory feedback could improve functional use of the prosthesis and reduce PLP. Importantly, we aimed to demonstrate that we could achieve these effects regardless of whether the amputation was traumatic or secondary to diabetic peripheral neuropathy, which substantially increases the pool of people that might benefit from these devices.

In three people with below-knee amputation (Table 1), we implanted commercially available SCS leads in the thoracolumbar epidural space to stimulate the lateral lumbar spinal cord. We identified electrode contacts that evoked sensation experienced on the missing foot and performed psychophysical assessments to characterize those sensations. We developed a closed-loop system (Figure 1) where SCS was modulated by pressure signals wirelessly recorded from an insole in the shoe under the prosthetic limb. Using this system to deliver real-time somatosensory feedback, we assessed balance and gait, as well as changes in PLP over the duration of the multi-week implantation period. Our results indicate that lumbar SCS is a promising intervention to restore sensations, improve function, and reduce PLP in lower limb amputees.

**Table 1.** Study participant demographics and amputation data.

| Subject | Age Range | Gender | Ambulation Level | Years since amputation | Side of amputation | Nature of amputation | Implant Duration (days) |
| --- | --- | --- | --- | --- | --- | --- | --- |
| 1 | 56-60 | M | Limited community | 3 | Left | Diabetic | 28 |
| 2 | 56-60 | M | Active | 7 | Left | Traumatic | 28 |
| 3 | 61-65 | W | Limited community | 5 | Left | Diabetic | 84 |

**Figure 1:**
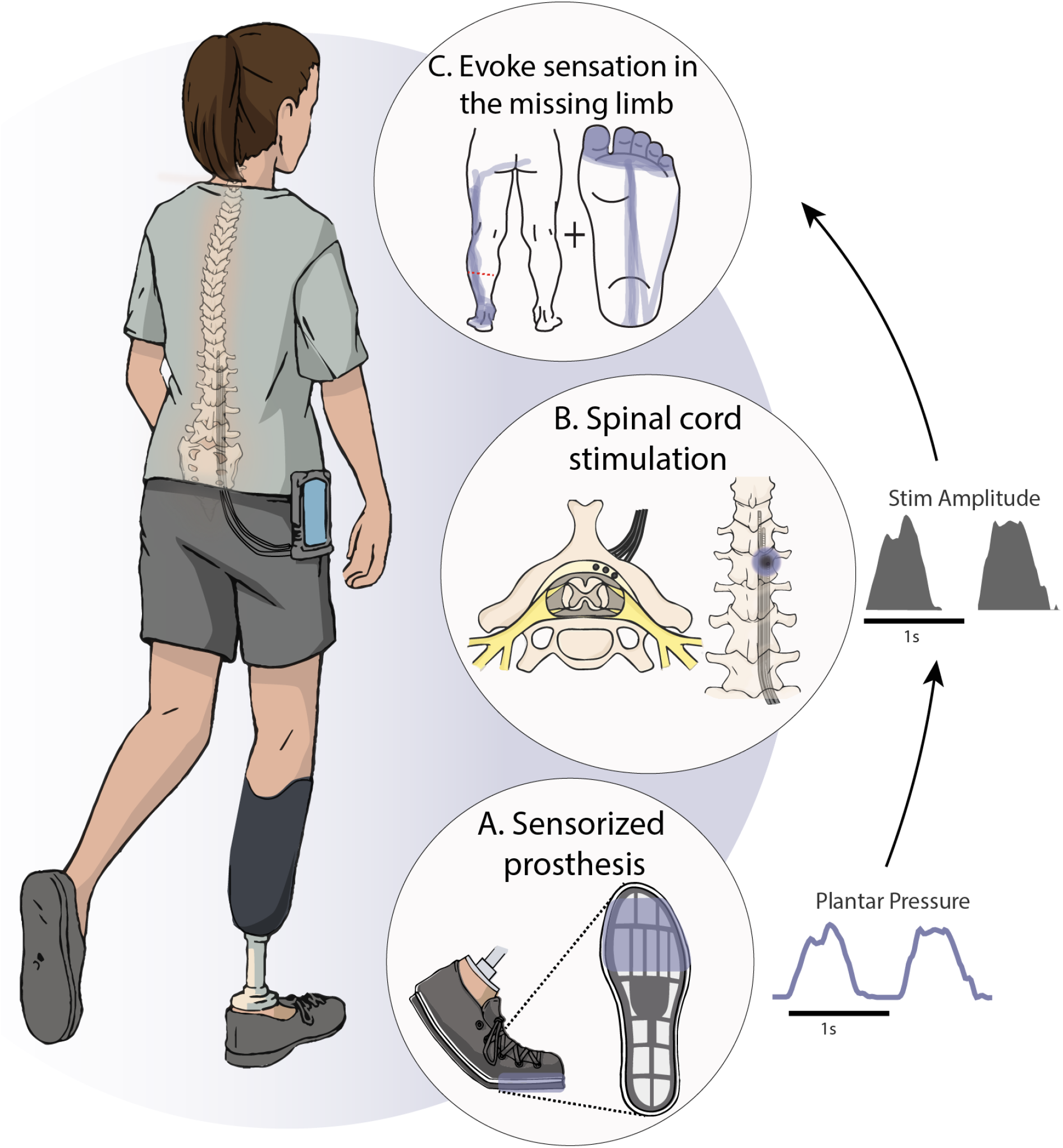
Schematic of the closed-loop SCS system used in this study. Electrical stimulation was delivered to the spinal cord via two or three 8- or 16-contact leads implanted percutaneously near the lateral lumbosacral spinal cord. The leads were tunneled through the skin and connected to an external stimulation system. (A) A sensorized insole was inserted into the shoe to measure pressure under the prosthetic foot, (B) the signals from this insole were used to modulate stimulation amplitude for SCS electrodes implanted in the lateral thoracolumbar epidural space, and (C) the stimulation evoked sensation that appeared to emannate from the missing limb. The purple line shows the location of evoked sensation in one subject and the red dotted line represents the end of the residual limb for this subject with left trans-tibial amputation.

### Spinal Cord Stimulation Evokes Sensations in the Missing Foot

The first goal of this study was to characterize the location and perceptual qualities of sensations evoked by lumbosacral SCS. To map the location of evoked sensations, we delivered 1-sec long stimulation trains, and asked the subjects to draw the location of the perceived sensations on a graphic representation of the foot and legs. For all three subjects, SCS evoked sensations in the missing limb, including the toes and heel (Figure 2A). The sensations were absent during the first 1-2 weeks of the study and appeared gradually thereafter (Figure 2C and Extended Data Figure 1). The sensations in the missing limb were always accompanied by sensations in the residual limb, and higher stimulation amplitudes were required to evoke sensations in the missing limb than in the residual limb alone (Extended Data Figure 2). The rostral-caudal arrangement of the electrodes across different levels of the spinal cord elicited sensations that corresponded to the dermatomal distribution^21^ (Figure 2B and Extended Data Figure 3).

**Figure 2:**
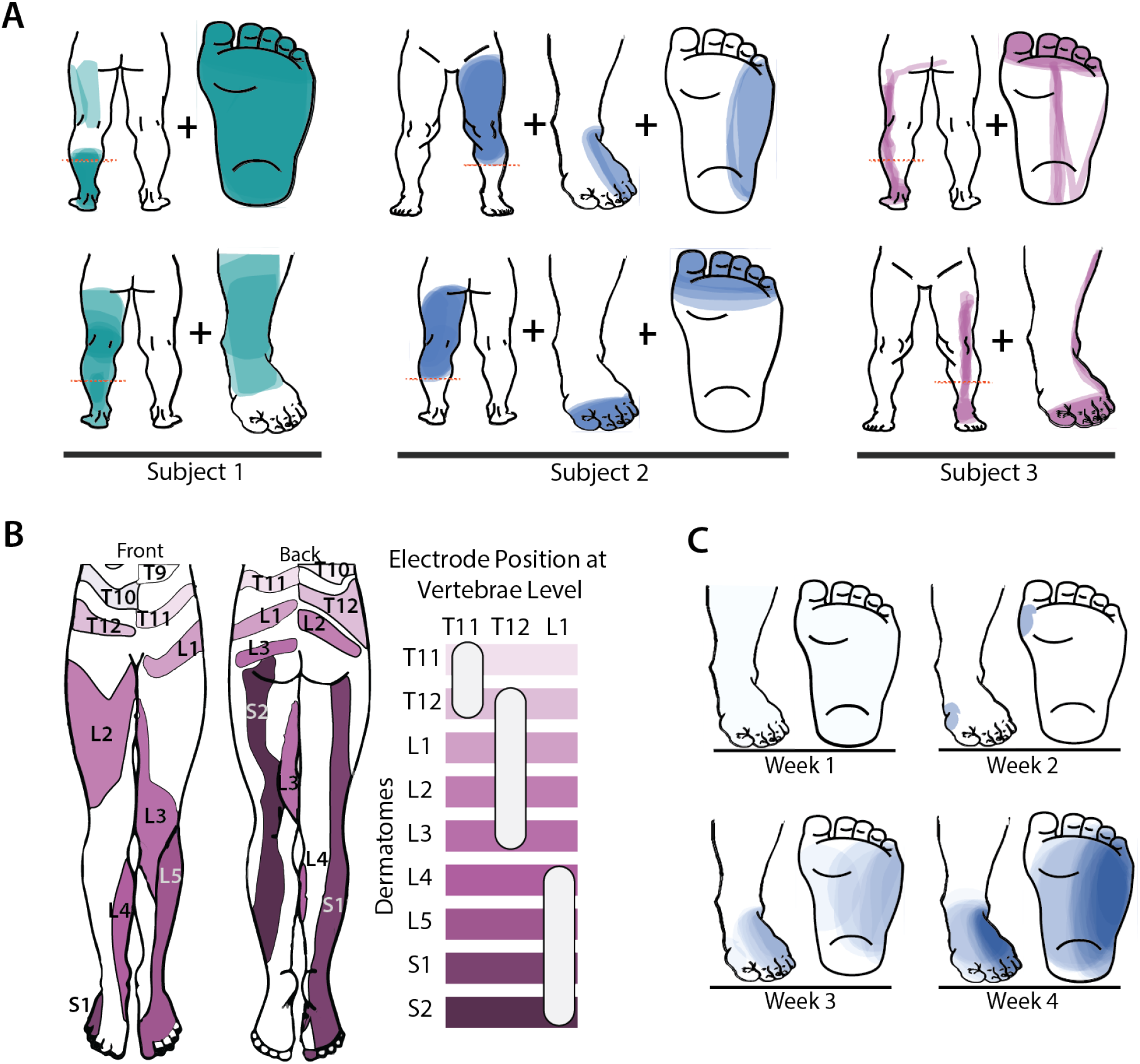
SCS evokes percepts in the missing limb. (A) Examples of percepts evoked in the missing and residual limb from one session for each subject. Two different sensations (corresponding to stimulation through two different electrodes) are shown for each subject (top and bottom). The red dashed line indicates the level of the amputation. The colored area represents the location of the perceived sensation, with darker colors representing more frequent reports of sensation at that location across trials, normalized within each subject. (B) Dermatome activation by electrodes located at different vertebrae levels for Subject 3. Left: Expected dermatomal innervation in the leg, adapted from Melzak et al.^21^ Right: Horizontal bars indicate different dermatomes and the white ovals indicate the approximate electrode position that evoked sensations in those dermatomes, with respect to vertebrae level. (C) Rate of occurence of sensations in the missing limb across weeks from one electrode in Subject 2. Darker shades indicate more frequent reports of evoked sensations in the foot.

The subjects also reported the perceived quality of the sensations using a list of descriptors compiled from previous literature^22^. For analytical purposes, we grouped these descriptors as sensations that subjects might experience commonly in their daily life (naturalistic) or rare, less familiar sensations (paresthetic). All subjects reported a combination of natural and paresthetic descriptors in different proportions (Extended Data Figure 4).

### Sensory magnitude can be systematically manipulated by varying stimulation amplitude

A key step in designing a sensory prosthesis is to assess the dependence of the sensation on stimulation parameters. With this in mind, we first established the stimulation intensity required to evoke a conscious percept. To this end, we had the subjects perform a detection task in a two-alternative forced choice paradigm. In brief, a 1-sec stimulation train at one of 5 to 10 amplitudes, determined in preliminary experiments to be peri-liminal, was presented in one of two visually cued stimulus intervals, and the subject’s task was to report which interval contained the stimulus. Each stimulus was presented at least 4 times and we tallied the proportion of times the subject correctly identified the interval containing the stimulus for each amplitude (Figure 3A). The detection threshold was the amplitude (estimated from the fitted logistic psychometric function) at which the subject would correctly identify the stimulus interval 75% of the time. Detection thresholds varied across electrodes and subjects from 0.6 to 4 mA but there were no large and systematic differences across subjects (Figure 3B).

**Figure 3:**
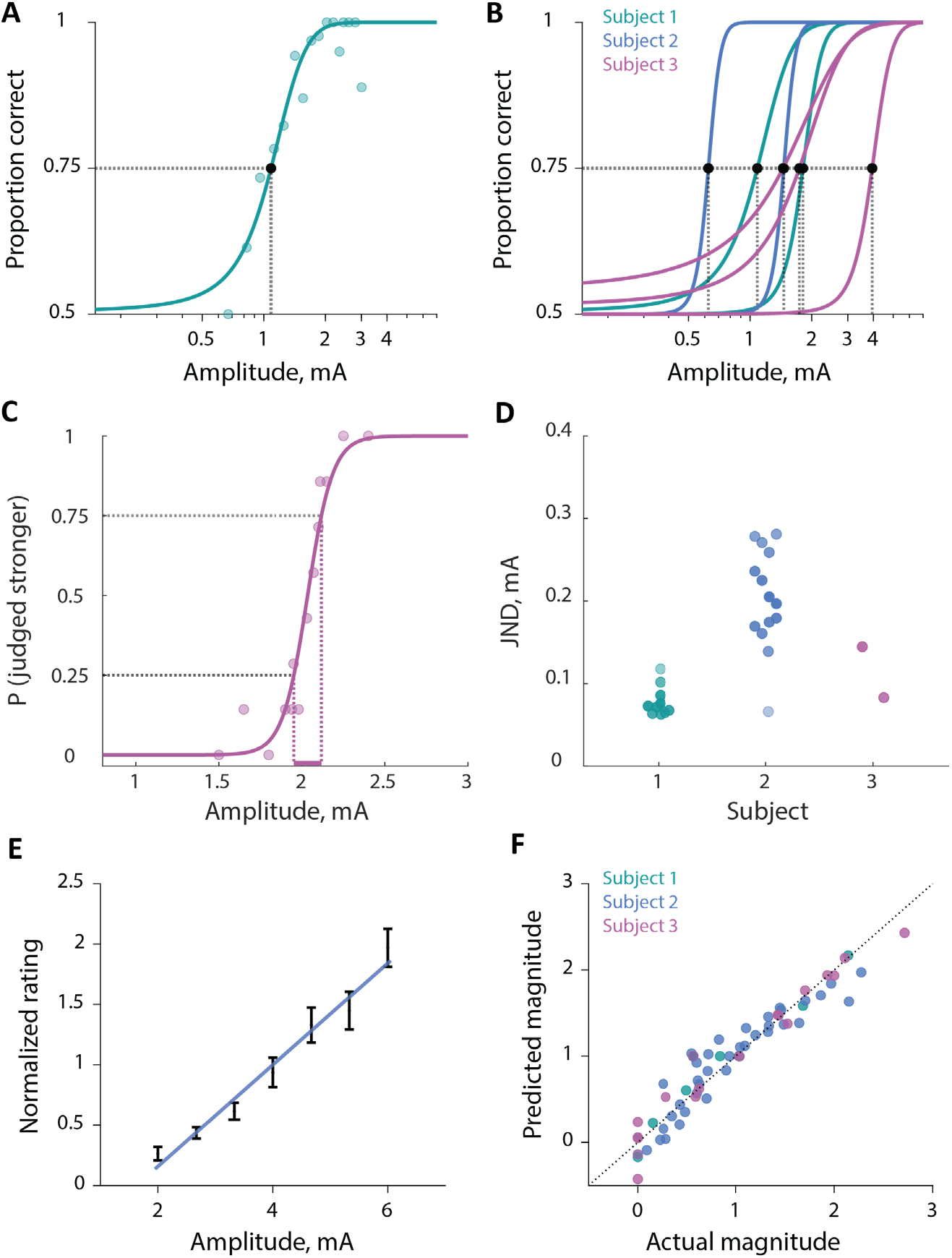
Psychophysical assesments of evoked sensations. (A) Performance of Subject 1 on the detection task for one electrode, showing the proportion of times the stimulus interval was correctly selected as a function stimulus amplitude. The bold line shows a cumulative-normal curve fit to the data. (B) Psychometric functions for the subset of electrodes (N=7 across all subjects) in which psychophysical assessment of detection threshold was assessed, color-coded by subject. The dashed lines indicate the detection threshold for each electrode. (C) Performance of Subject 3 on the amplitude discrimination task with a standard amplitude of 2 mA for one electrode. The bold line shows the fitted psychometric function and the dashed lines indicate the range used to compute the just noticeable difference (JND) (in this case 0.08 mA), given by half the distance between the dotted lines. (D) Distribution of JNDs across the three subjects on a subset of electrodes (N=10 sessions from 1 electrode for Subject 1, N=14 sessions from 3 electrodes for Subject 2, and N=2 sessions from 2 electrodes for Subject 3). (E) Average normalized magnitude ratings as a function of stimulus amplitude for one electrode for Subject 2. The bold line indicates the linear fit to the data. The error bar denotes the standard deviation across repeated presentations of the same stimulus. (F) Scatter plot showing predicted magnitude estimated by a linear model for each electrode (N=3 sessions from 1 electrode for Subject 1, N=11 sessions from 4 electrodes for Subject 2 and N=4 sessions from 3 electrodes for Subject 3) and subject versus the actual stimulation magnitude. Points show average magnitude for each presented amplitude for each electrode, color-coded by subject. The dashed line represents the unity line.

Next, we measured the subjects’ sensitivity to changes in stimulation amplitude. To achieve this, we had them perform an amplitude discrimination task. On each trial, the subject was presented with two stimuli: (1) a standard whose amplitude was fixed within the block, and (2) a comparison, whose value varied from trial to trial. After both presentations, the subject reported which of the two felt stronger (Figure 3C). For each electrode and subject, we fitted a logistic psychometric function and computed the just noticeable difference (JND), the change in amplitude required for the subject to correctly identify the more intense stimulus 75% of the time. JNDs varied from 0.05 to 0.3 mA across subjects and electrodes (Figure 3D, Extended Data Figure 5). The range of JNDs overlapped across subjects, though Subject 2 tended to have higher JNDs than the other two subjects.

Finally, we wished to explicitly measure the relationship between stimulation amplitude and perceived magnitude. To this end, we delivered stimuli that spanned a range of intensities and had the subject report how intense the stimulus felt with the following instructions: (1) If they did not feel the stimulus; they ascribed to it a rating of 0; (2) If one stimulus felt twice as strong as another, it was to be ascribed a number that was twice as high (other examples were also provided); (3) They could use any scale they wanted, and were encouraged to use decimals, if necessary. Perceived magnitude increased nearly linearly with stimulation amplitude for all subjects and electrodes (R^2^ of the linear regression for Subject 1 was 0.978, Subject 2 was 0.854 and Subject 3 was 0.952, Figure 3E,F), as has been previously found with stimulation of the peripheral nerves^23–26^ and of the somatosensory cortex^27^. Because of this linear relationship between the perceived intensity and stimulation amplitude, we used linear modulation of stimulation amplitude in subsequent experiments assessing functional outcomes (see below).

### Spinal cord stimulation improves functional use of a prosthesis

The second goal of this study was to demonstrate that restored somatosensation can improve functional use of a prosthetic limb. To restore somatosensation during functional tasks, such as standing and walking, we placed a wireless pressure-sensing insole (Moticon Insole 3, Munich, Germany) under the prosthetic foot and used the output from that insole to control stimulation in real-time. In Subjects 2 and 3, we selected an SCS electrode that reliably evoked sensation on the plantar surface of the missing foot and used the pressure signal from the same location under the prosthetic foot to control stimulation amplitude (Figure 1A). Due to time constraints, we could not perform these experiments in Subject 1. We used clinical measures of balance and gait to compare postural stability with and without restored somatosensory feedback.

#### Spinal cord stimulation improves standing balance

To assess standing balance with and without sensory feedback, we used the Sensory Organization Test (SOT), a clinical outcome measure that quantifies reliance on visual, vestibular, or somatosensory feedback to maintain balance control. The SOT requires the subject to maintain balance (Figure 4A) despite erroneous visual information from a visual surround that can sway and/or erroneous somatosensory information from a support surface that can also sway. To characterize reliance on vision, vestibular sense, and somatosensory feedback, the SOT comprises six different conditions, which each condition obscuring different combinations of the relevant sensory feedback. With somatosensory feedback restored via SCS, both Subjects 2 and 3 achieved improvements in SOT scores (Figure 4C), with greater improvements in the most challenging conditions (platform sway with eyes closed and platform sway with visual surround sway, Extended Data Figure 6). Notably, both participants experienced at least one “fall” without stimulation, but neither subject fell with stimulation (Figure 4C). A “fall” denotes a failure to complete the trial due to taking a step, falling in the harness, or grabbing the walls for support. Performance was slightly worse with stimulation during the least challenging conditions (i.e., no visual or support surface sway) with eyes open and eyes closed (Extended Data Figure 6), although this difference was negligible (i.e., smaller than the minimum clinically important difference). In Subject 2, we implemented a sham stimulation condition, in which stimulation evoked sensation only in the residual limb and not in the missing foot. In this case, we saw decreased performance from baseline for conditions 2 (static platform with eyes closed), 5, (platform sway with eyes closed), and 6 (platform sway with visual surround sway) and the decrease for condition 6 was larger than a minimum clinically important difference (Extended Data Figure 6). Biomechanical analyses of center of gravity traces (COG, Figure 4D) revealed that both participants exhibited statistically significant decreases in sway area (indicating greater stability) with eyes closed condition and an unstable support surface during stimulation (Subject 2 decreased by 19.34 cm^2^, Subject 3 decreased by 39.04 cm^2^, p<0.001, Figure 4D).

**Figure 4.**
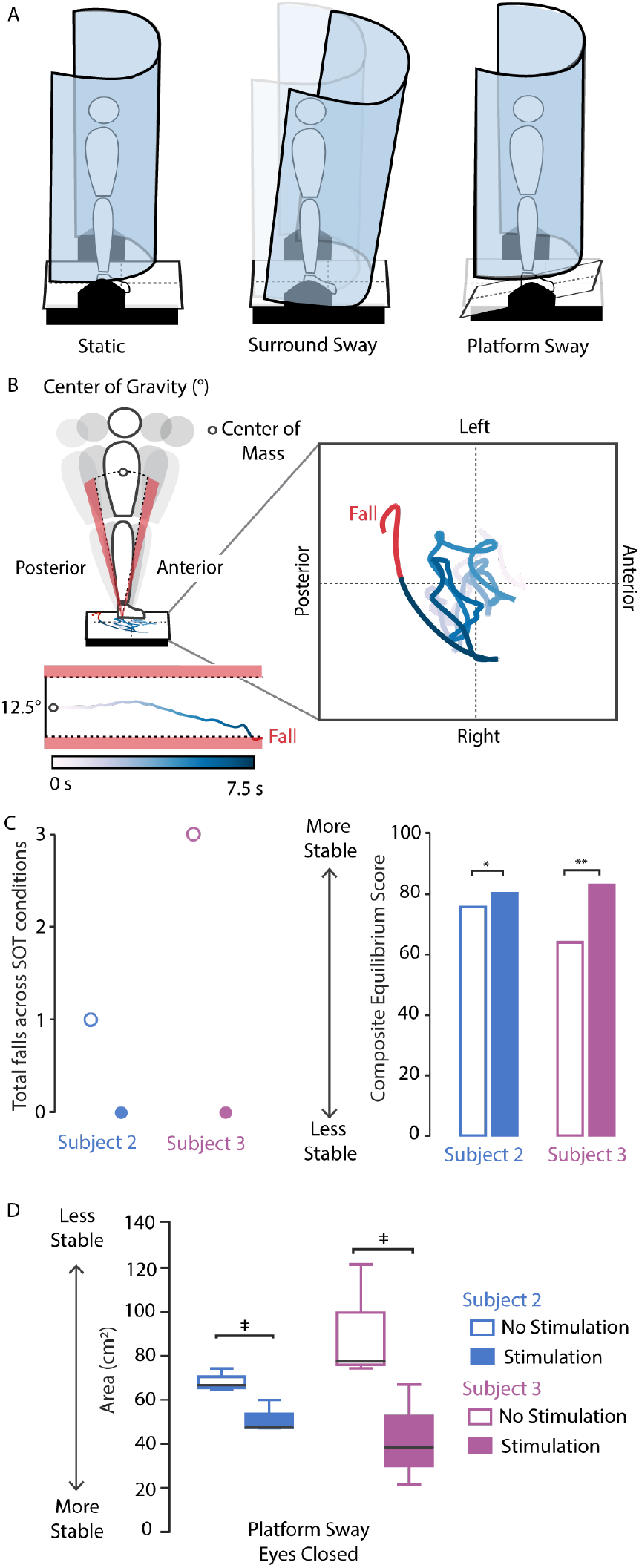
Closed-loop sensory feedback improves postural stability. (A) The Sensory Organization Test (SOT) comprises six conditions, defined by whether the visual surround (middle) and/or platform (right) are swaying while subjects have their eyes open or closed. (B) For analysis, the center of gravity (COG) is a projection of the pressure trace onto the force plate to indicate their center of mass (COM) movement throughout a trial. The equilibrium score is an indication of how well subjects maintain their COM within a normative 12.5° limit of anteroposterior sway. Beyond these limits, a fall can occur (red). (C) Falls occurred only during conditions without stimulation for both subjects (left) and both subjects exhibited an improvement in composite equilibrium score (right). This improvement was above the minimum dectectable change for Subject 2 (*) and above the threshold for a clinically meaningful difference in Subject 3 (**). (D) Both subjects showed a statistically significant decrease in sway area, indicating greater stability, with stimulation (‡).

#### Spinal cord stimulation improves gait stability

To assess stability during gait, participants performed the Functional Gait Assessment (FGA), a clinical measure of dynamic balance, commonly applied to detect reliance on visual and somatosensory systems for maintaining balance during walking^28,29^. This task consists of ten items, including walking with eyes closed, walking with a narrow base of support, and walking over obstacles. Restored somatosensation led to a clinically meaningful improvement (>4 points) in FGA score for Subject 3 but not Subject 2 (Figure 5B). Notably, Subject 2 demonstrated baseline performance 3.9 points below age-matched able-bodied controls, whereas the baseline score for Subject 3 was 13.5 points below age-matched normative data^28^.

**Figure 5:**
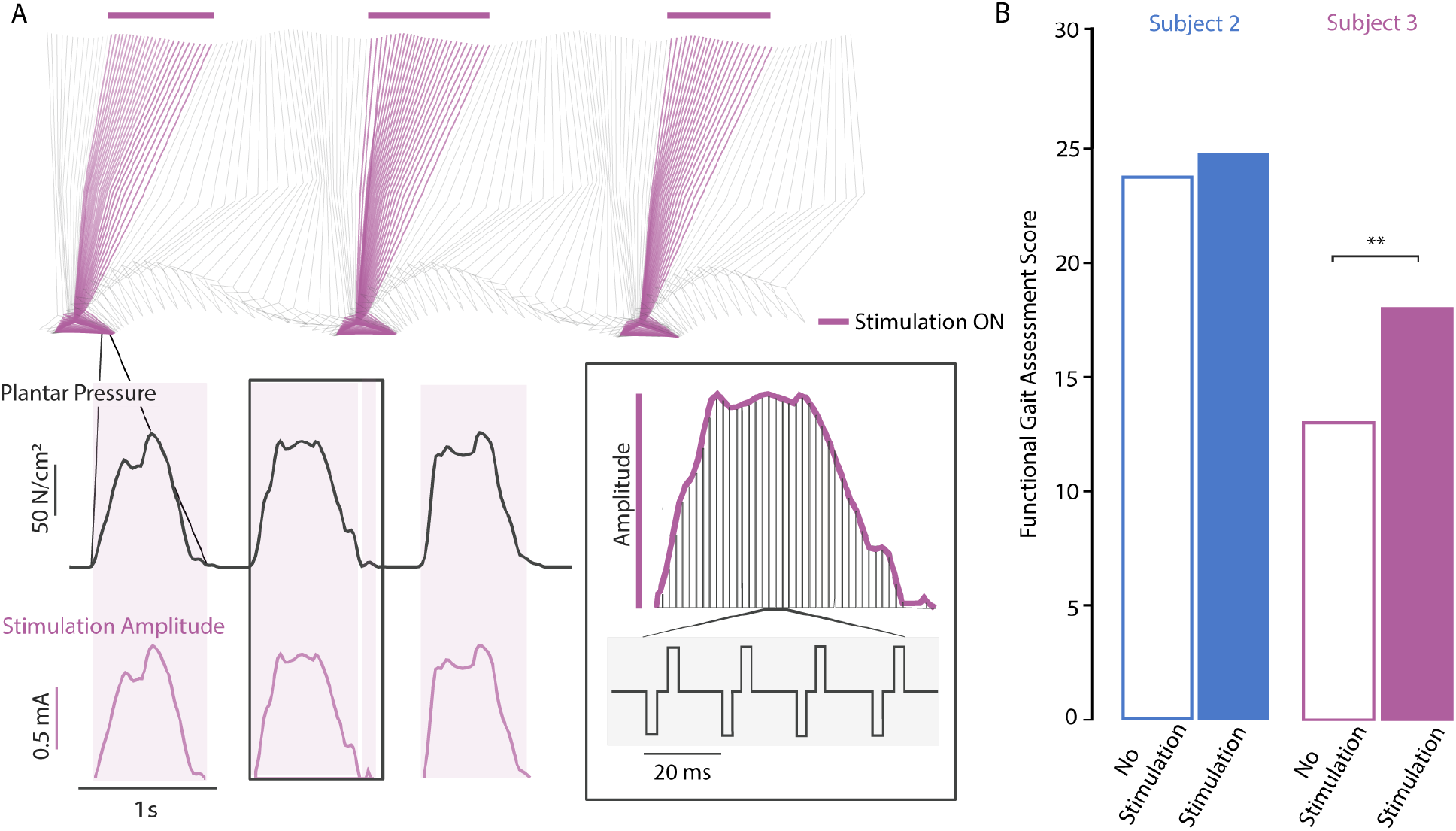
Closed-loop sensory feedback improves gait stability. (A) Example of amplitude modulation with plantar pressure throughout the gait cycle. Stimulation was triggered above a threshold for the metatarsals (purple shading) and either maintained at a constant amplitude (for Subject 2) or amplitude was modulated linearly with pressure signals (for Subject 3). (B) The Functional Gait Assessment (FGA) increased in both subjects, and increased beyond the minimum detectable change (MDC >4 points, **) for Subject 3, who had a lower baseline score.

### Spinal cord stimulation reduces phantom limb pain

To assess the impact of stimulation on PLP, we examined subjects’ reports of their current pain level on a visual-analog scale. Subjects were instructed to report the pain level perceived specifically in the missing limb for this assessment. As the study progressed, we observed a clinically meaningful decrease in PLP score (defined as a 50% reduction from the baseline pain score) for Subjects 1 and 3 (Fig. 6). While the PLP score for Subject 2 also decreased to 0.73 from 1.2 points, this improvement is considered sub-clinical because it is less than 1 point. For both Subjects 1 and 3, the first clinically meaningful decrease in PLP coincided with the emergence of electrically evoked sensations in the missing limb (i.e. week 3 and week 2, respectively). For Subject 3, experiments were suspended over a one-week holiday (week 11), at which time pain scores increased sharply (3.65 times greater than week 10), consistent with the hypothesis that SCS relieves PLP.

**Figure 6:**
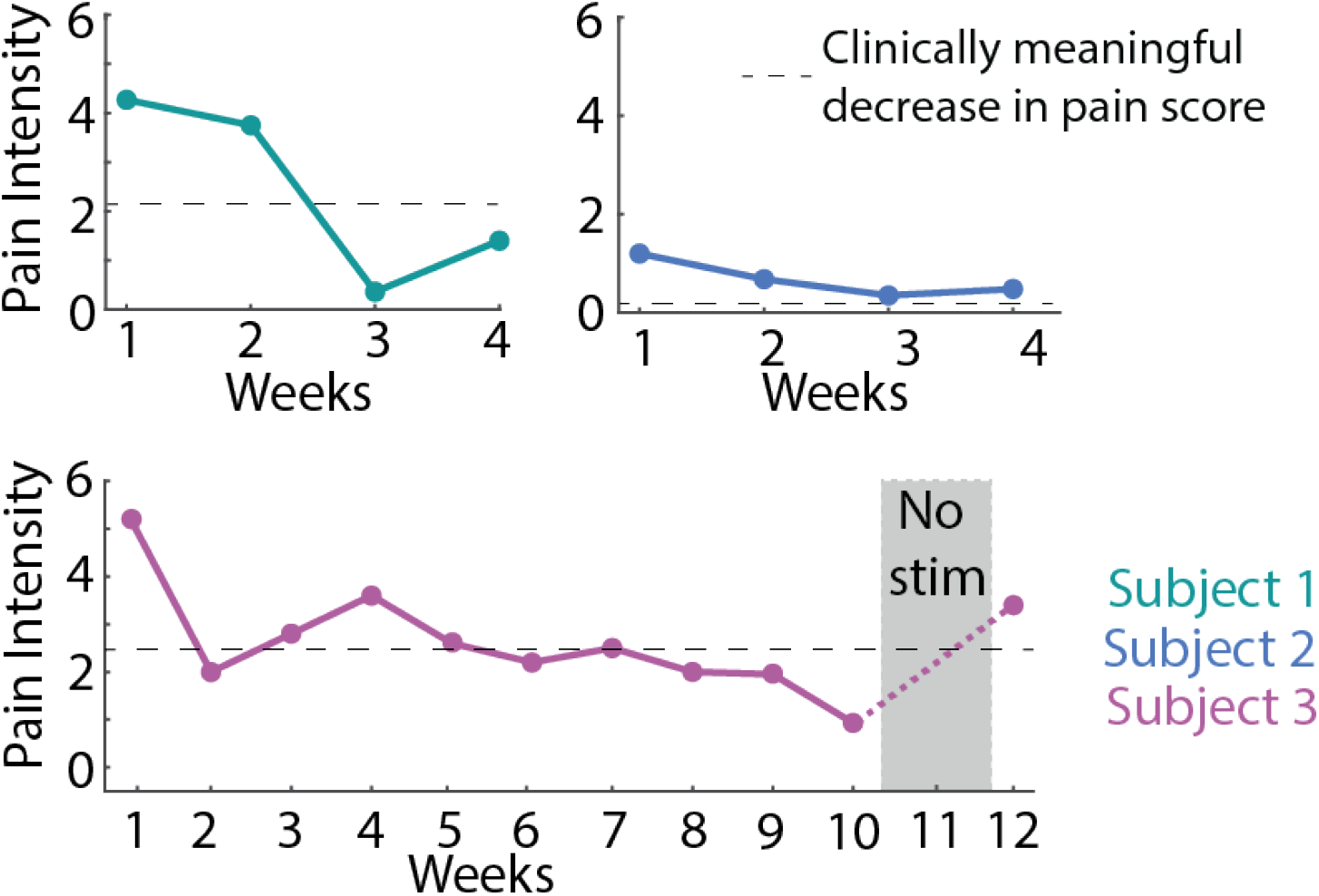
Spinal cord stimulation reduces phantom limb pain. Phantom limb pain intensity as reported weekly on a visual analog scale for Subject 1 and 2 (top row, implanted for 4 weeks) and Subject 3 (bottom row, implanted for 12 weeks). The dashed line indicates a clinically meaningful decrease in the pain score. For Subject 3, no experiments were conducted during week 11 (marked with a gray box).

We also conducted the McGill Pain Questionnaire^30^ once per week, to characterize the sensory and affective dimensions of the subjects’ pain. This questionnaire provides a holistic measure of a patients pain experience and can be used to infer overall patient well-being. All subjects were instructed to rate their pain in the missing limb over the most recent week of the study (Extended Data Figure 7). We observed a clinically meaningful decrease in the McGill Pain Questionnaire scores (defined as a >5-point decrease) for Subject 1 (28 points) and Subject 2 (10 points) across the 4-week implant phase. For Subject 3, across the 12-week implant, there was a reduction in the pain scores until week 8 (15 points) followed by an increase from week 9 onwards, including a 24 point increase that coincided with the break in testing during week 11. However, Subject 3 anecdotally reported a substantial reduction in PLP episodes.

## DISCUSSION

In this study, we demonstrate that lateral lumbosacral SCS evokes sensations in the missing limb in people with transtibial amputation, and that this restored somatosensation can improve balance control, gait stability, and reduce PLP. Importantly, we showed these effects across a range of subjects, including people with amputations that occurred long before enrollment in the study (up to 7 years), a subject with traumatic amputation and two others with diabetic peripheral neuropathy, and people with different levels of mobility. Critically, the implantable electrodes used in this study were commercially available devices that are currently implanted in more than 50,000 people each year for the treatment of pain^18^. The devices were implanted via a percutaneous approach in an outpatient surgical procedure, and future development and translation of our approach can leverage the vast infrastructure of clinicians and surgical techniques that already exist for SCS. While translation will still require substantial technical and clinical development, this study demonstrates the feasibility of using SCS to restore somatosensation from the missing foot with the potential to improve quality of life for people with lower-limb amputation.

In all subjects, we found that multiple SCS electrodes evoked sensations in the missing limb, and each subject reported more than one sensation in different locations on the missing limb. However, we also found that the sensations evoked in the missing limb always co-occurred with sensations in the residual limb. Subject 1 and 2 reported simultaneous sensations in distinct areas of the residual and missing limb, whereas Subject 3 reported contiguous sensations spanning the residual and missing limb. In a previous study that focused on people with upper-limb amputation, we observed similar coincidence of SCS-evoked sensations in the residual and missing limb in only two out of four subjects, and the sensations on the residual limb tended to be more focal in the arm than in the leg^20^. This difference may reflect anatomical differences between the cervical and lumbar regions of the spinal cord. Indeed, sensory neurons enter the cervical spinal cord at a shallow angle, nearly perpendicular to the rostrocaudal axis, whereas they travel parallel to the rostrocaudal axis for several spinal segments before entering the lumbar cord^31^. Accordingly, afferents that innervate multiple regions of the legs are more densely packed in the lumbar region than in the cervical region. Delivering charge in the epidural space using the large commercially available SCS electrodes likely recruits more sensory afferents in the lumbar cord, increasing the likelihood of activating neurons projecting from both missing and residual limb. Moving forward, designing SCS leads with smaller and more densely packed electrodes may allow us to achieve more selective stimulation and consequently, more focal sensations in the missing limb.

We also found that subjects did not report sensations in the missing limb until the second or third week of the study. During the first 1-2 weeks, a majority of reported sensations were diffuse and limited to the residual limb. Following this period, subjects consistently reported sensations in the missing limb. Other studies using peripheral nerve stimulation to evoke sensation in the missing foot have also reported that the incidence of sensations in the missing limb increases with time^32^. This delayed emergence of sensations in the missing foot stands in contrast to our previous study, in which subjects frequently reported sensations in the missing upper limb during intraoperative testing of cervical SCS^20^.

We found that the magnitude of electrically-evoked sensations can be systematically manipulated by modulating the stimulation amplitude, as has been previously reported across a variety of stimulation modalities and neural targets^20,24^. JNDs ranged from around 0.05 to 0.3 mA across subjects and leads (mean = 0.15 mA). The JNDs were independent of the reference amplitude (Extended Data Figure 5), violating Weber’s law, which states that the JND should be directly proportional to reference amplitude [ref]. This is similar to results reported for both peripheral nerve stimulation^24^ and for intracortical microstimulation^33^. Together, these results suggest that approximately 20 discriminable steps can be achieved from threshold (typically less than 2 mA) to maximum amplitude (4-6 mA). The dynamic range of SCS-based tactile feedback is thus comparable to or wider than its counterparts in the peripheral nerve^24^.

One of the goals of this study was to demonstrate that restored somatosensation could improve standing balance and gait stability. We found that SCS-evoked somatosensory feedback improved standing balance, particularly in the more challenging conditions (in which visual and somatosensory feedback were altered), consistent with previous results using peripheral nerve stimulation to restore sensation in the foot^10^. Furthermore, the reduction of falls in the SOT with stimulation constitutes a critical improvement in balance control. Note that these substantial and clinically meaningful improvements in balance were observed even though evoked sensations extended from the missing limb onto the residual limb. While evoking focal sensations in the missing limb is likely to further improve balance, our results suggest even non-focal sensations projecting from both the missing and residual limb may be sufficient to improve function.

During gait, we saw a clinically meaningful improvement in the Functional Gait Assessment for Subject 3. Notably, Subject 3 had a lower baseline score than Subject 2, allowing the possibility for greater improvements with stimulation. Because this clinical assessment serves as a relatively crude measure of dynamic balance control during ambulation, it may not be sensitive to changes with sensory feedback and, like many other clinical measures, is subject to ceiling effects. Additionally, we have recently demonstrated that spatiotemporal analysis of level walking is insensitive to large differences in somatosensory ability across individuals with an amputation and is similarly unlikely to be able to reveal subtle improvements with restored sensation^34^. These findings indicate that, while we see improvements with stimulation, we should identify more sensitive and more challenging outcome measures to detect improvements in function with restored somatosensory feedback. Additionally, evaluating fall risk itself over a longer time period will be critical in future studies to demonstrate the clinical importance of restored somatosensation after amputation.

In addition to functional improvements, we also found evidence that stimulation reduced PLP. Subjects 1 and 3 showed a clinically significant decrease in PLP during the week in which they first reported experiencing evoked sensations in the missing limb. This observation is similar to the gradual decrease in PLP reported in other studies with lower-limb amputees and suggests neuroplastic changes in the brain may follow evoked sensations in the missing limb^8^. Subject 3 also reported that PLP increased when testing was paused for a week. The recurrence of PLP aligns with anecdotal evidence from traditional SCS studies which report that the wash-in and wash-out period of SCS can be 3-7 days^35^. Our observations build on growing evidence that somatosensory neuroprosthetic systems are associated with a decrease in PLP^7,11–14^.

Several important limitations remain to be addressed in future studies. First, the subject pool in this study was small and heterogenous, including two people with diabetic neuropathy and substantial mobility limitations and a third person with a traumatic amputation and a high degree of active mobility. Further, the study was not blinded to either the subject or the investigators. It is possible that subjects were motivated to better performance when they knew that we were delivering stimulation. However, the use of the sham stimulation in Subject 2 suggests that providing sensory feedback in the residual limb was insufficient to improve balance, and instead sensation in the missing limb was important for this effect. While we demonstrated initial feasibility of our approach, larger randomized controlled trials will be critical for demonstrating that SCS can improve function and reduce PLP after lower-limb amputation. Second, the intervention in this study involved a percutaneously implanted device tested over 29 or 90 days in a laboratory setting. Future studies should include a fully implanted system, including an implantable pulse generator wirelessly communicating with external sensors on the prosthesis, as well as long-term testing of performance in real-world settings. Third, the stimulation delivered during this study involved simple, constant frequency trains in which amplitude was modulated based on pressure signals from an insole under the prosthetic foot. More complex trains of stimuli, such as biomimetic patterns that more closely match the naturalistic patterns of activity in somatosensory afferents, may produce more naturalistic sensations, yielding greater functional gains and possibly stronger pain relief^36–39^.

This study marks an important step towards clinical translation of somatosensory neuroprosthetics for people with lower-limb amputation. We demonstrated that SCS delivered via commercially available electrodes, implanted through a common clinical procedure, could evoke sensations in the missing foot in people with transtibial amputation. Importantly, this includes two people with amputations related to diabetic peripheral neuropathy. We also demonstrated in two subjects that these sensations, when controlled by a wireless insole in the shoe, improved balance control and gait stability. Finally, we measured decreases in phantom limb pain in all three subjects during their participation in the study. Building on these promising results, we believe that SCS may be a clinically viable approach to restore sensation and improve quality of life for people with transtibial amputation.

## METHODS

### Subjects

Three subjects with transtibial amputation (Table 1) were recruited for this study. Two subjects had diabetes and peripheral neuropathy associated with the amputations, while one subject had a traumatic amputation. All subjects were active users of a non-motorized prosthetic limb before beginning the study. The two subjects with diabetes (Subjects 1 and 3) were limited community ambulators and the subject with traumatic amputation was an active ambulator (exceeding community ambulation skills, Subject 2). The study was approved by the University of Pittsburgh Institutional Review Board and the extended duration implant in Subject 3 was performed under an Investigational Device Exemption from the U.S. Food and Drug Administration. Both studies (i.e. 29-day and 90-day implant periods) are registered at ClinicalTrials.gov (NCT03027947 and NCT04547582). Subjects provided informed consent prior to participation.

#### Inclusion/Exclusion Criteria

Subjects 21-70 years old were included in the study if they had a unilateral transtibial amputation, and were not excluded for partial amputation (e.g. one or more toes) on the contralateral limb. Subjects were at least 6 months post-amputation at the time of SCS lead implantation, with no serious comorbidities that could increase risk of participation. Woman who were pregnant or breast feeding, people taking anticoagulant drugs, and people with implanted metal not cleared for MRI or implanted medical devices such as pacemakers, defibrillators, and infusion pumps were excluded. Subjects were also excluded from the 90-day implant study if they had a hemoglobin A1c level above 8.0 mg/dl, because of the increased infection risk associated with this condition.

### Electrode Implant

SCS leads were implanted percutaneously via a minimally invasive procedure, under local and/or twilight anesthesia. Subjects were in the prone position while leads were inserted using into the dorsal epidural space using a 14-guage 4-inch epidural Tuohy needle, and the leads were steered posterior laterally using a stylet under live fluoroscopic guidance. The connector from each lead was externalized so that we could connect it to an external stimulator. We performed intraoperative stimulation and subjects verbally reported the location of evoked sensations so we could iteratively adjust the placement of the leads to evoke sensations in the missing limb or as close to the end of the residual limb as possible. In Subject 1, two 16-contact leads (Infinion, Boston Scientific) were implanted near the T12-L2 vertebral levels and a third 16-contact lead was inserted through the sacral hiatus to target the cauda equina. The third lead did not produce useful sensations in the missing limb, so this type of insertion was not repeated in subsequent subjects. In Subject 2, two 16-contact leads (Infinion, Boston Scientific) were inserted near the T12-L2 vertebral levels. In Subject 3, three 8-contact leads (Octrode, Abbott Medical) were inserted near the T12-L2 levels. Lead migration was monitored by comparing intraoperative fluoroscopic images to weekly X-rays for the first 4 weeks and then bi-monthly X-rays for the following weeks in Subject 3. In Subject 1, leads were anchored with sutures to the superficial layers of skin at the exit sites, and all three leads demonstrated substantial caudal migration across weeks during the implant. Therefore, to better stabilize the electrode placements, in Subjects 2 and 3 the leads were anchored to subcutaneous fascia via a small incision. With this anchoring procedure, we saw minimal lead migration across weeks. At the end of the study, a similar procedure was performed to remove all leads from the body.

### Mapping Evoked Sensations

To map the location of evoked sensations, we stimulated each electrode contact using a 1-sec long charge-balanced bi-phasic pulse train. We stimulated with amplitudes from 0.5 to 6 mA and with frequencies from 1 to 1000 Hz. The pulse width was fixed at 200 μs and the interphase interval was set to 60 μs. Stimulation was delivered using a custom-built circuit board and three 32-channel stimulators (Nano 2+Stim; Ripple, Inc)^20^.

After each stimulation train, the subject reported the location and quality of the sensation using our previously developed touchscreen interface^40^. The quality of the sensation was described using a predefined list of descriptors developed specifically for characterizing sensations evoked by electrical stimulation^22^, including mechanical, movement, tingle, and temperature sensations. For analytical purposes, we grouped these descriptors as sensations that subjects might experience commonly in their daily life (naturalistic) or rare, less familiar sensations (paresthetic). In total, 13 descriptors were used for naturalistic modalities (pulsing, pressure, touch, sharp, tap, urge to move, vibration, flutter, itch, tickle, prick, cool, and warm) and 5 descriptors were used for paresthetic modalities (electric current, tingle, buzz, shock, and numb).

### Psychophysical analysis: Detection threshold estimation

We used a two-alternative forced choice task where the subjects were presented with two visually cued 1-sec long blocks with a variable delay period: one with stimulation and one without stimulation, assigned randomly. The subjects were instructed to select the block in which they felt a sensation. The stimulus amplitudes were centered around the rough detection threshold we observed from the mapping trials on that day. Overall, stimulus amplitudes ranged from 0.5 to 6 mA and each amplitude was repeated 4-10 times. The stimulation frequency remained constant at 50 Hz for all trains. For each stimulus amplitude, we calculated the number of times the subject responded correctly (accuracy rate). For electrodes with densely sampled stimulation amplitudes (that differ by less than 0.1mA), the values were re-binned with 0.1 mA steps and the amplitudes that were in the same interval bin were replaced by their mean. A logistic curve constrained between 0.5 and 1 was fit to the accuracy rate for each subject and electrode, and the stimulus amplitude corresponding to 75% accuracy rate was selected as the detection threshold. Electrodes with insufficient repetitions per condition (<5) or poor logistic fit (goodness of fit of the model is insignificant at 10%) were excluded from analysis. Overall, we tested two electrodes for Subject 1, two electrodes for Subject 2 and three electrodes for Subject 3.

### Psychophysical analysis: Just noticeable differences

We used a similar two-alternative forced choice task to determine the just noticeable difference for the evoked sensations (i.e. the minimum detectable change in stimulation amplitude). On each trial, the subject was presented with two stimuli: (1) a standard whose amplitude was fixed within the set, and (2) a comparison, whose value varied from trial to trial. The subject was asked to report which of the two stimuli felt more intense. Standard amplitudes across different sets ranged from 1-3.5 mA for Subject 1, 1.2-4.55 mA for Subject 2, and 2-4.74 mA for Subject 3. Comparison amplitudes ranged from 50 to 150% of the standard amplitude in that set. The frequency and pulse width of both standard and comparison stimuli remained constant (50 Hz, 0.2 ms). In each set, each stimulus pair was presented at least 5 times, and both the order of stimuli within the pair and the order of the pairs were varied pseudo randomly.

A logistic function was fit to the percentage of times the comparison interval was selected by the subject to obtain psychometric curves for each standard amplitude. Then, the just noticeable difference (JND) was calculated as the change in amplitude that led to 75% accuracy according to the psychometric curve. We tested for 10 sessions from 1 electrode for Subject 1, 14 sessions from 3 electrodes for Subject 2, and 2 sessions from 2 electrodes for Subject 3. Sets with poor psychometric fits (goodness of fit of the model is insignificant at 10%) were omitted from the analysis.

### Psychophysical analysis: Perceived stimulation intensity

To understand the relationship between the stimulus amplitude and the perceived intensity of the sensation, we conducted a magnitude estimation experiment. On each trial, a 1-sec long pulse train was delivered, and the subject was asked to state a number whose magnitude corresponded to the magnitude of the evoked sensation. Subjects were instructed to use their own scale including decimals. If a stimulus was imperceptible, it was ascribed to the number 0. If one stimulus felt twice as intense as another, it was given a number that was twice as large. The tested amplitudes ranged from 0.5 to 6 mA and were restricted to amplitudes above detection threshold for each channel. The maximum amplitude delivered was below the pain/discomfort threshold for each subject. Each test amplitude was presented at least 5 times.

Ratings of sensation magnitude were normalized by the mean rating of their respective set and linear regression was fit to the observed data for each channel separately. The residuals of regression models were tested for normality with Kolmogorov-Smirnov test to justify linear fit. We tested for 3 sessions from 1 electrode for Subject 1, 11 sessions from 4 electrodes for Subject 2, and 4 sessions from 3 electrodes for Subject 3.

### Closed-loop stimulation for functional tasks

To use these evoked sensations for real-time feedback in a functional task, such as gait or balance, data from wireless plantar pressure sensing insoles (Moticon Insole 3, Munich, Germany) was used to trigger stimulation of the spinal cord in real-time. For Subjects 2 and 3, one electrode that evoked sensations in the missing limb was selected to provide real-time feedback of plantar pressure. In both participants, the evoked sensations were in the toes and metatarsals (Figure 1). Because of time constraints, Subject 1 did not participate in this portion of the study. Plantar pressure above a minimum threshold triggered stimulation in the same region as the mapped sensation. Subject 2 experienced sustained quadriceps contractions for stimuli above 2.5 mA. Because of the small range of stimulation amplitudes available between threshold and these contractions (2.25-2.5 mA), for Subject 2 we utilized constant amplitude stimulation for, in which stimulation turned on when insole pressure was above a threshold and turned off when below that threshold. For Subject 3, plantar pressure linearly modulated stimulation amplitude; as she put more weight on her metatarsals, she felt a more intense sensation (Figure 5A). Stimulation frequency (50 Hz Subject 1, 90 Hz Subject 2) and pulse width (200 μs for both subjects) were kept constant and amplitude was updated every 20 ms.

### Functional assessments

The Sensory Organization Test (SOT) was used to determine changes in balance ability using the NeuroCom Equitest system (Figure 3A). The SOT is a clinical measure of reliance on visual, vestibular, and somatosensory systems for balance using six conditions where either the surround or platform sway. Three 20-second trials were completed per condition. The SOT was completed pre-implant without stimulation and repeated later with stimulation. Center of pressure (COP) traces were recorded from the support surface at 100 Hz, filtered with a low-pass fourth-order Butterworth filter, and analyzed for biomechanical and clinical measures of posturography. Standard posturography measures were calculated, including excursion, sway velocity, 95% confidence interval ellipse of sway area, sample, and approximate entropy^41^. The primary clinical outcome measure for each condition is the equilibrium score, a measure of the participant’s ability to stay within a normative 12.5° of anteroposterior sway (Figure 3B).

During walking, the Functional Gait Assessment (FGA) and kinematics of walking on a level surface were evaluated. The FGA is a 10-item test of dynamic balance, including challenging items like walking with eyes closed, walking with a narrow base of support, and walking backwards^28^. Each item is scored on a scale from 0 to 3 points, where 3 indicates no impairment and 0 indicates an inability to complete the task. Gait kinematics were recorded with a 16-camera OptiTrack motion analysis system (Natural Point, OR, USA). Sixteen reflective markers were placed on anatomical landmarks according to the OptiTrack “Conventional Lower Body” model. Motion capture data were collected at 100 Hz and filtered using a 4th order low-pass Butterworth filter at 12 Hz. Participants were instructed to walk at their self-selected speed across a 6-meter walkway. For Subject 2, 14 trials of walking without stimulation and with stimulation were analyzed. For Subject 3, 28 trials without stimulation and with stimulation were compared.

For clinical measures, published standards for clinically meaningful difference (CMD) or minimum detectable change (MDC) were used to compare baseline and stimulation trials^42–44^. For biomechanical measures of balance and gait, comparisons between outcomes were performed using permutation testing, a non-parametric method often used for smaller sample sizes^45^. We completed 10,000 permutations of both baseline and with-stimulation groups and the difference in means of biomechanical data across trials was determined. The p-value in permutation testing is the count of permutations in which the observed difference in means is above the actual difference in means. An alpha of 0.05 was used for all statistical analyses.

### Phantom Limb Pain

To quantify phantom limb pain, we asked the subjects to report their current pain level on a visual analog scale (VAS) at the beginning of each testing day. The scale ranged from 0 to 10 where 0 indicated no pain and 10 indicated the worst pain imaginable. VAS scores were averaged over each week. Typically, a 50% decrease (and at least a 1-point decrease) in VAS score is considered clinically meaningful.

Subjects also completed the McGill Pain Questionnaire (MPQ) once per week to describe their pain over the previous week. The MPQ is a multi-dimensional survey of the affective, evaluative, and other experiences of pain and requires the subject to select from ranked lists of descriptor words (such as dull, sore, hurting, aching, heavy) about their pain. Subjects also select a value ranging 0-5 to describe the intensity of their pain. The total score from this instrument is intended to reflect both the intensity and the disruptive nature of pain, and a 5-point decrease is considered clinically meaningful.

## Data Availability

All data produced in the present study are available upon reasonable request to the authors.

## Extended Data

**Extended Data Figure 1:**
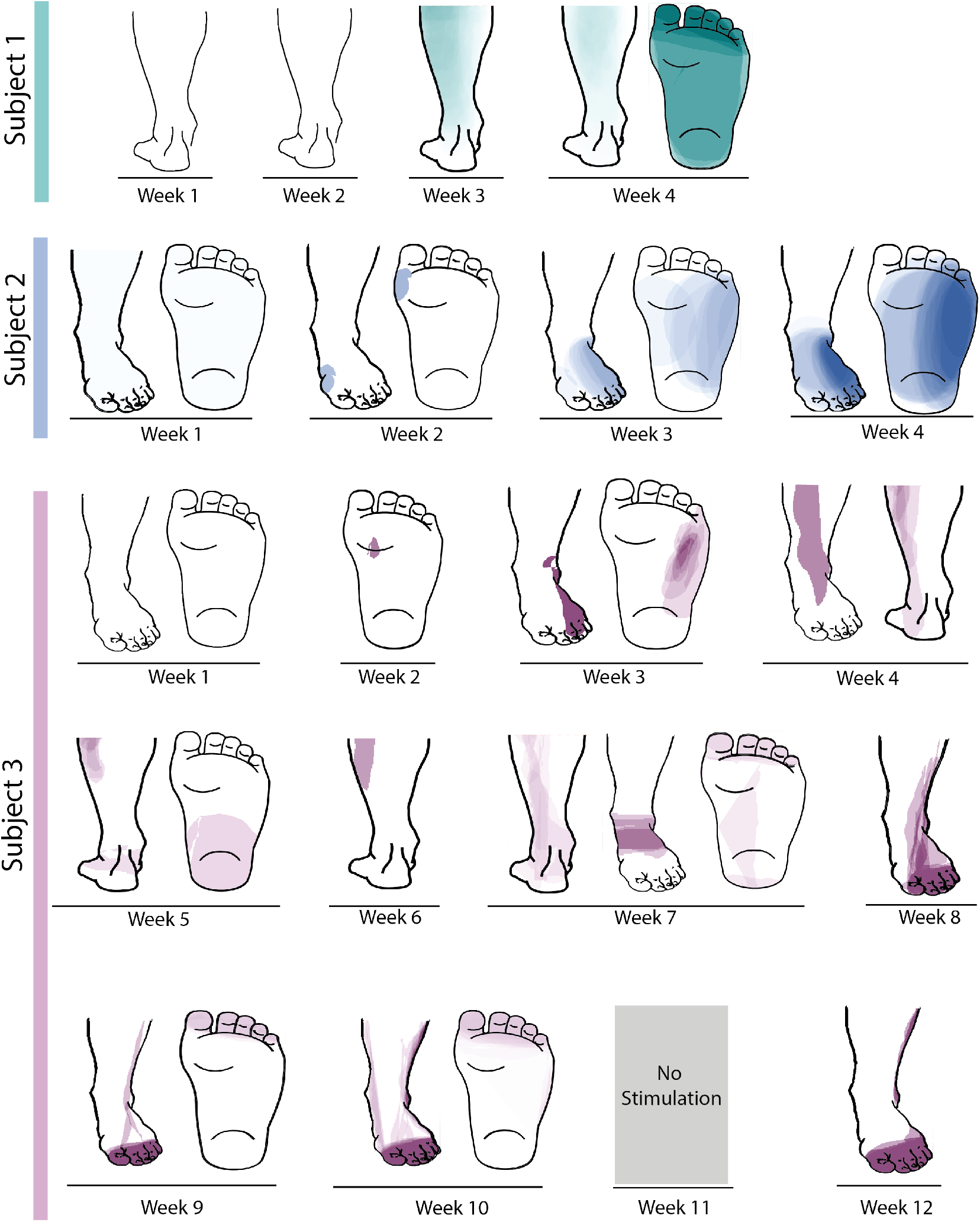
Heatmaps showing the rate of occurrence of sensations in the missing limb across weeks. Darker shades indicate higher rate of occurrence of sensations in that location. No testing was done on week 11 for Subject 3.

**Extended Data Figure 2:**
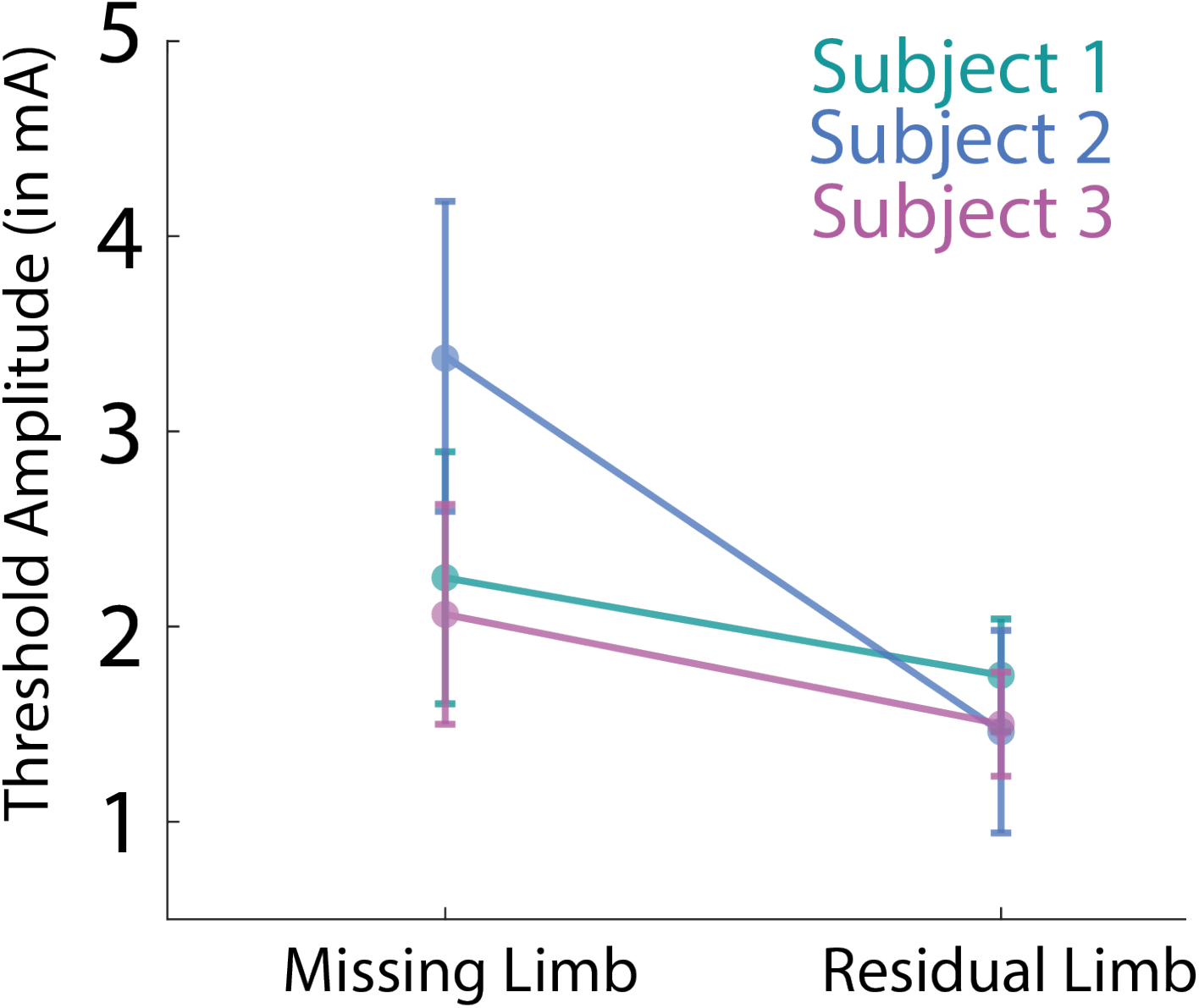
Comparison of the threshold amplitude that evoked sensation in missing limb (with co-activation in the residual limb) and the threshold amplitude that evoked sensation only in the residual limb. The threshold amplitude for each testing day was determined by increasing the stimulation amplitude in 0.5 or 1 mA steps and asking the subjects to report the location where they perceived the evoked sensation. Error bars show the standard deviation across multiple days.

**Extended Data Figure 3:**
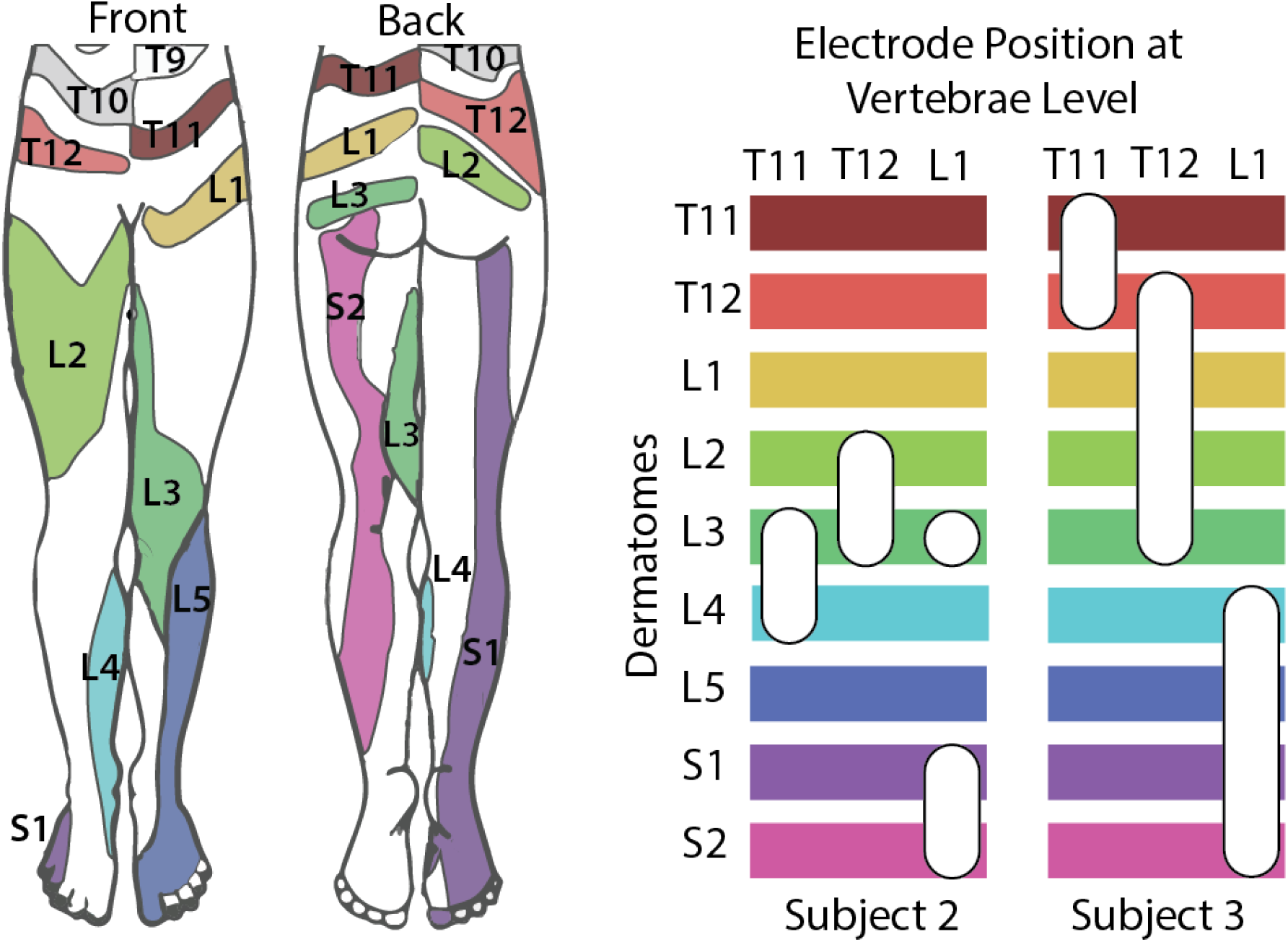
Dermatomal activation by electrodes located at different vertebrae levels for Subject 2 and Subject 3. The left image shows the expected dermatomal innervation in the leg. In the right, the horizontal bars indicate different dermatomes^21^ and the white ovals indicate the approximate electrode position that evoked sensations in that dermatome with respect to the vertebrae level. Subject 1 had substantial lead migration across weeks, making it challenging to precisely define the location of the electrodes with respect to vertebrae levels, so we have not included those results.

**Extended Data Figure 4:**
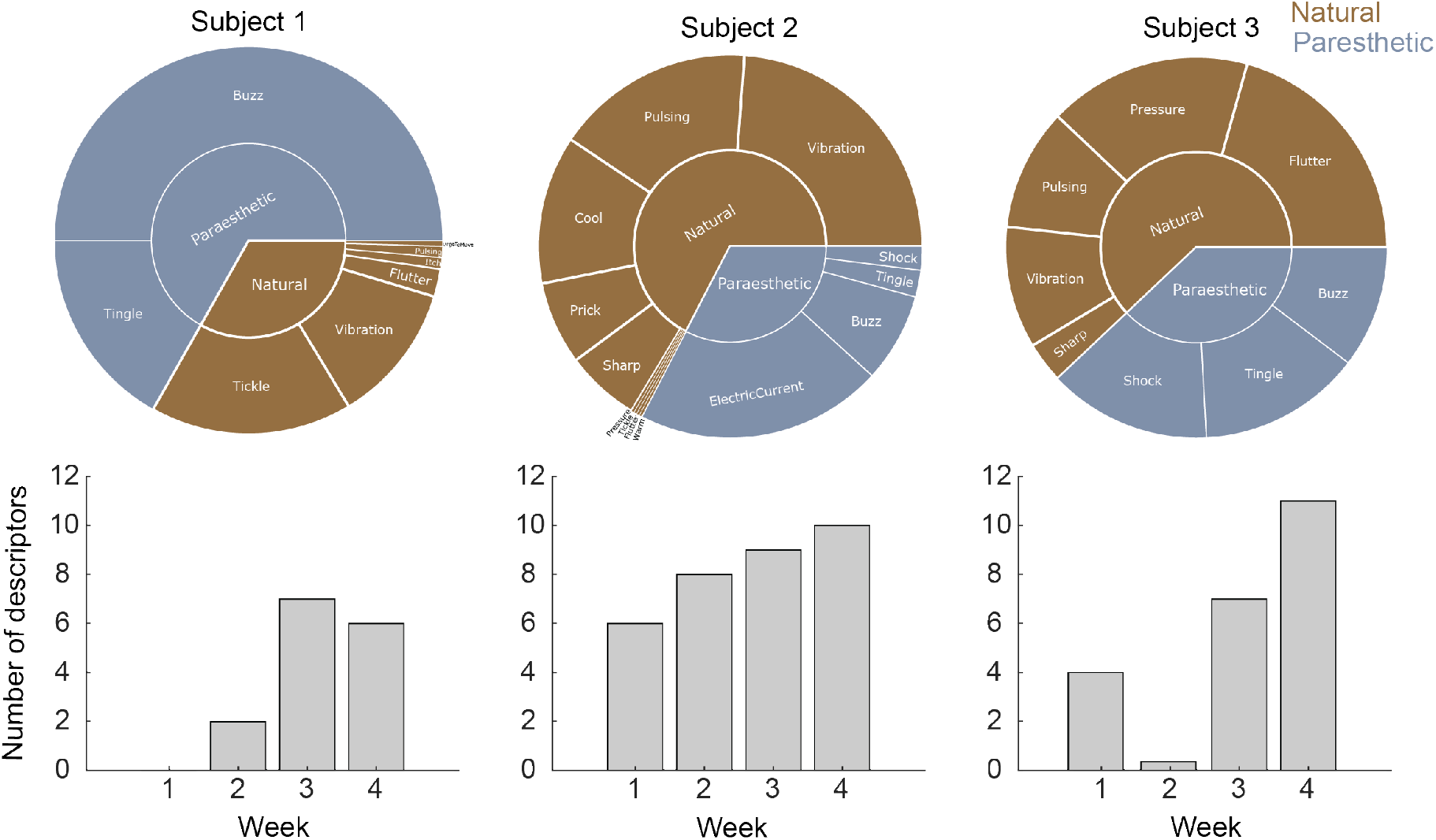
Percept quality of evoked sensations in the missing limb. The subjects were given a list of 13 natural descriptors and 5 paresthetic descriptors to describe the quality of the sensation. The top panel shows the frequency of each descriptor for the two evoked sensations for each subject shown in Figure 2a. For all reported sensations, we stimulated each electrode with a 1-sec long pulse train. The bottom panel shows the total number of descriptors used to describe the sensations each week.

**Extended Data Figure 5:**
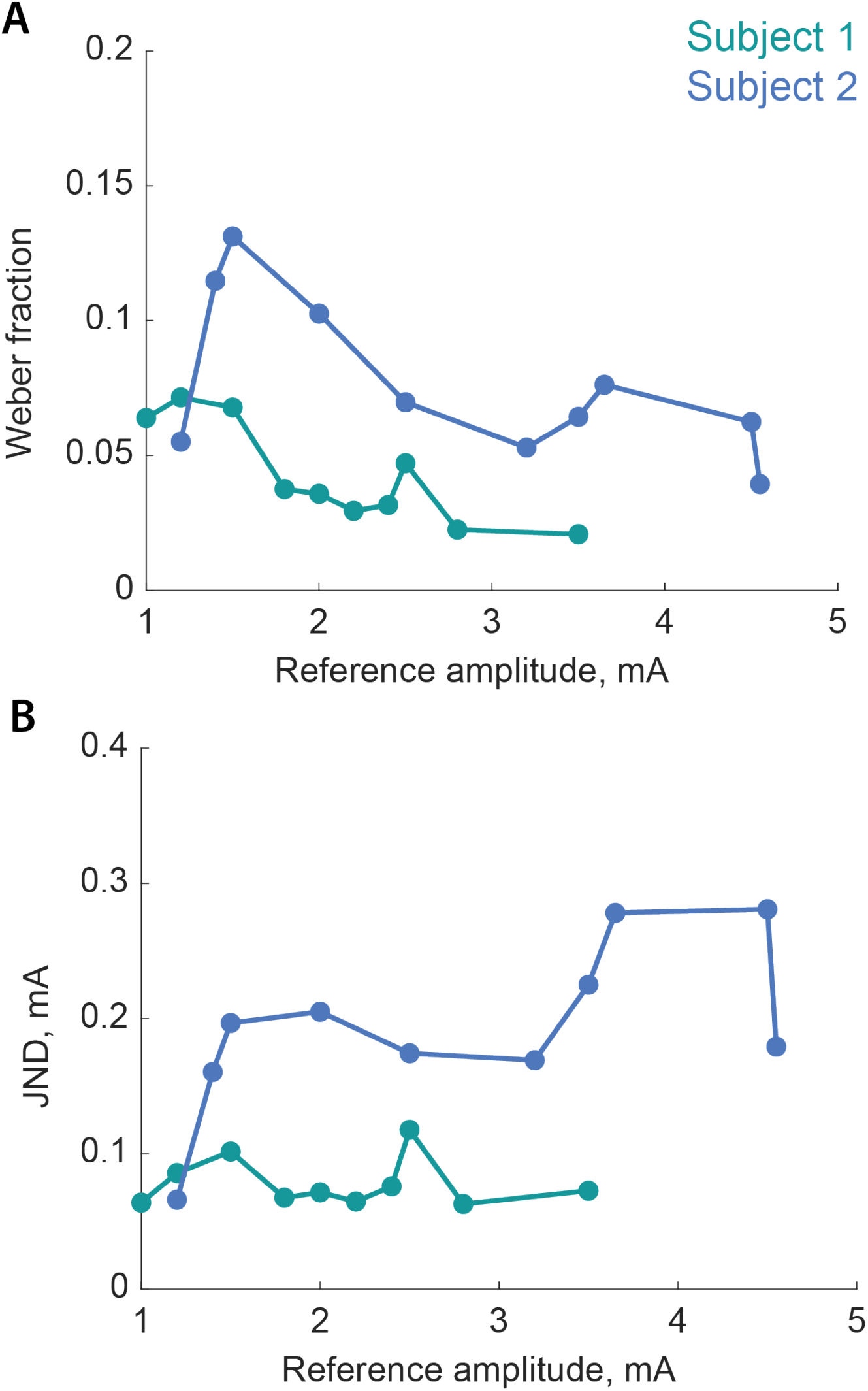
Additional results from psychophysical discrimination assessment. (A) Variation of Weber fraction for different electrodes in Subject 1 and 2 as a function of the reference amplitude in the discrimination task. (B) Variation of JND for the same electrodes in Subjects 1 and 2 as a function of the reference amplitude. Subject 3 was discarded from these analyses due to insufficient data points.

**Extended Data Figure 6:**
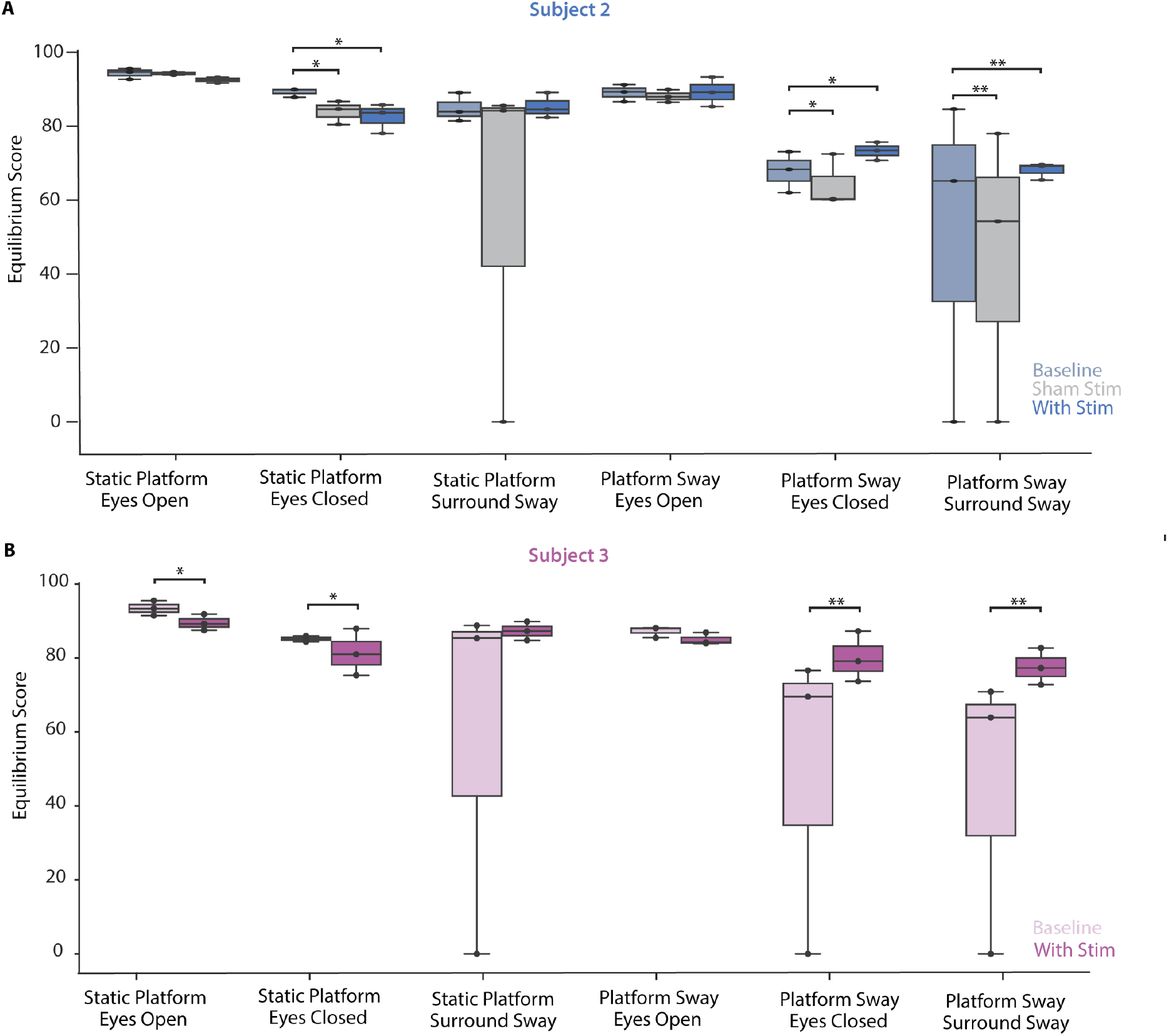
Full results of Sensory Organization Test (SOT). (A) Subject 2 performed the SOT without stimulation (light blue) with sham stimulation (i.e., stimulation **i**n the residual limb only, gray) and with stimulation (stimulation in the prosthetic foot, dark blue). Sham stimulation substantially decreased performance for three of six conditions (with greater than minimum detectable change [MDC, 3.98]), suggesting that stimulation on the residual limb alone was not sufficient to improve performance. Subject 3 performed the SOT without stimulation (light magenta) and with stimulation (dark magenta). Both Subject 2 and Subject 3 exhibited improved performance on conditions with platform sway and eyes closed (+5.12 Subject 2, +9.60 Subject 3) and with visual surround sway (+4.04 Subject 2, +13.39 Subject 3). Both subjects, however, exhibited decreased performance with stimulation during static standing with eyes closed (−6.25 Subject 2, -4.32 Subject 3). Additionally, Subject 3 had worse performance on static standing with eyes open with stimulation (−4.13). Change in median values reported. * represents a MDC, ** represents a clinically meaningful difference (>8.0).

**Extended Data Figure 7:**
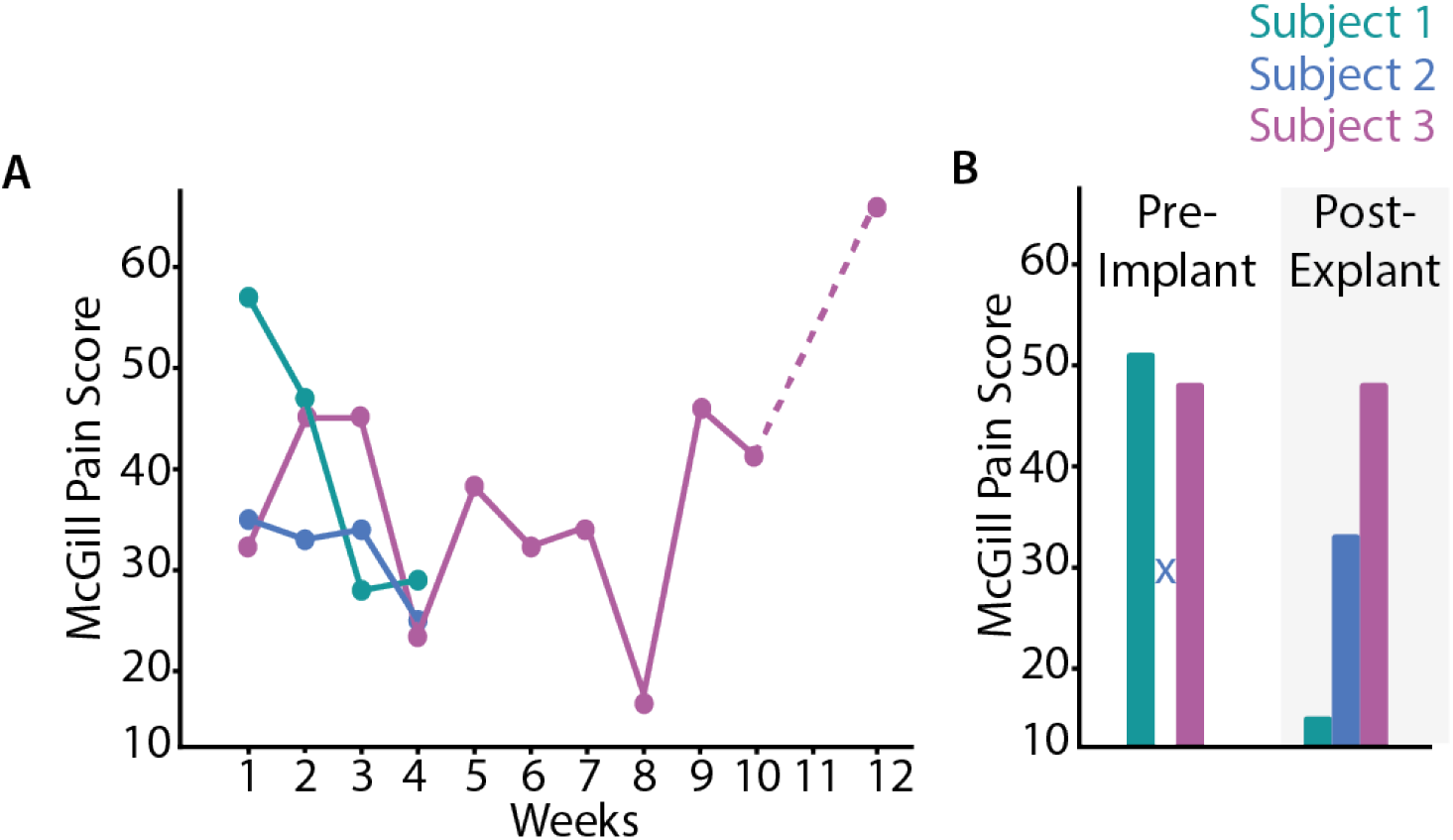
(A) Weekly McGill Pain Questionnaire results. (B) McGill Pain Questionnaire score before the implant and 1-month post-explant. The pre-implant score for Subject 2 was not recorded and we did not perform testing on week 11 for Subject 3 (indicated by the dashed line).

## References

1. Sexton, A. T. & Fleming, L. L. Lower extremity amputations. in Medical Management of the Surgical Patient: A Textbook of Perioperative Medicine (eds. Walker, H. K., Lubin, M. F., Spell, N. O., Smith, R. B. & Dodson, T. F.) 741–743 (Cambridge University Press, 2006). doi:DOI: 10.1017/CBO9780511544590.119.

2. Viseux, F. J. F. The sensory role of the sole of the foot: Review and update on clinical perspectives. Neurophysiol. Clin. 50, 55–68 (2020).

3. Petersen, B. A., Nanivadekar, A. C., Chandrasekaran, S. & Fisher, L. E. Phantom limb pain: peripheral neuromodulatory and neuroprosthetic approaches to treatment. Muscle Nerve 59, 154–167 (2019).

4. Valle, G. et al. Mechanisms of neuro-robotic prosthesis operation in leg amputees. Sci. Adv. 7, eabd8354 (2021).

5. Petrini, F. M. et al. Enhancing functional abilities and cognitive integration of the lower limb prosthesis. Sci. Transl. Med. 11, eaav8939 (2019).

6. Charkhkar, H. et al. High-density peripheral nerve cuffs restore natural sensation to individuals with lower-limb amputations. J. Neural Eng. 15, 056002 (2018).

7. Petrini, F. M. et al. Sensory feedback restoration in leg amputees improves walking speed, metabolic cost and phantom pain. Nat. Med. 25, 1356–1363 (2019).

8. Raspopovic, S. et al. Restoring natural sensory feedback in real-time bidirectional hand prostheses. Sci. Transl. Med. 6, 222ra19 (2014).

9. Dhillon, G. S. & Horch, K. W. Direct neural sensory feedback and control of a prosthetic arm. IEEE Trans. Neural Syst. Rehabil. Eng. 13, 468–472 (2005).

10. Charkhkar, H., Christie, B. P. & Triolo, R. J. Sensory neuroprosthesis improves postural stability during Sensory Organization Test in lower-limb amputees. Sci. Rep. 10, 6984 (2020).

11. Ortiz-Catalan, M., Hakansson, B. & Branemark, R. An osseointegrated human-machine gateway for long-term sensory feedback and motor control of artificial limbs. Sci. Transl. Med. 6, 257re6–257re6 (2014).

12. Horch, K., Meek, S., Taylor, T. G. & Hutchinson, D. T. Object discrimination with an artificial hand using electrical stimulation of peripheral tactile and proprioceptive pathways with intrafascicular electrodes. IEEE Trans. Neural Syst. Rehabil. Eng. 19, 483–489 (2011).

13. Tan, D. W. et al. A neural interface provides long-term stable natural touch perception. Sci. Transl. Med. 6, 257ra138–257ra138 (2014).

14. Davis, T. S. et al. Restoring motor control and sensory feedback in people with upper extremity amputations using arrays of 96 microelectrodes implanted in the median and ulnar nerves. J. Neural Eng. 13, 036001 (2016).

15. Marasco, P. D., Schultz, A. E. & Kuiken, T. A. Sensory capacity of reinnervated skin after redirection of amputated upper limb nerves to the chest. Brain 132, 1441–1448 (2009).

16. Rossini, P. M. et al. Double nerve intraneural interface implant on a human amputee for robotic hand control. Clin. Neurophysiol. 121, 777–783 (2010).

17. Dillingham, T. R., Pezzin, L. E. & MacKenzie, E. J. B. T.-S. M. J. Limb amputation and limb deficiency: epidemiology and recent trends in the United States. 95, 875+ (2002).

18. Kumar, K. & Rizvi, S. Historical and Present State of Neuromodulation in Chronic Pain. Curr. Pain Headache Rep. 18, 387 (2014).

19. Fanciullo, G. J., Rose, R. J., Lunt, P. G., Whalen, P. K. & Ross, E. The State of Implantable Pain Therapies in the United States: A Nationwide Survey of Academic Teaching Programs. Anesth. Analg. 88, (1999).

20. Chandrasekaran, S. et al. Sensory restoration by epidural stimulation of the lateral spinal cord in upper-limb amputees. Elife 9, e54349 (2020).

21. Lee, M. W. L., McPhee, R. W. & Stringer, M. D. An evidence-based approach to human dermatomes. Clin. Anat. Off. J. Am. Assoc. Clin. Anat. Br. Assoc. Clin. Anat. 21, 363–373 (2008).

22. Kim, L. H., McLeod, R. S. & Kiss, Z. H. T. A new psychometric questionnaire for reporting of somatosensory percepts. J. Neural Eng. 15, 13002 (2018).

23. Raspopovic, S. et al. Restoring Natural Sensory Feedback in Real-Time Bidirectional Hand Prostheses. Sci. Transl. Med. 6, 222ra19–222ra19 (2014).

24. Graczyk, E. L. et al. The neural basis of perceived intensity in natural and artificial touch. Sci. Transl. Med. 8, 1–11 (2016).

25. Page, D. M. et al. Discriminability of multiple cutaneous and proprioceptive hand percepts evoked by intraneural stimulation with Utah slanted electrode arrays in human amputees. J. Neuroeng. Rehabil. 18, 12 (2021).

26. Mastinu, E. et al. Grip control and motor coordination with implanted and surface electrodes while grasping with an osseointegrated prosthetic hand. J. Neuroeng. Rehabil. 16, 49 (2019).

27. Flesher, S. N. et al. Intracortical microstimulation of human somatosensory cortex. Sci. Transl. Med. 8, 361ra141–361ra141 (2016).

28. Wrisley, D. M. & Kumar, N. A. Functional Gait Assessment: Concurrent, Discriminative, and Predictive Validity in Community-Dwelling Older Adults. Phys. Ther. 90, 761–773 (2010).

29. Koehler-McNicholas, S. R., Danzl, L., Cataldo, A. Y. & Oddsson, L. I. E. Neuromodulation to improve gait and balance function using a sensory neuroprosthesis in people who report insensate feet – A randomized control cross-over study. PLoS One 14, e0216212 (2019).

30. Melzack, R. The McGill Pain Questionnaire: major properties and scoring methods. Pain 1, 277–299 (1975).

31. Cho, T. A. Spinal cord functional anatomy. Contin. Lifelong Learn. Neurol. 21, 13–35 (2015).

32. Charkhkar, H. et al. High-density peripheral nerve cuffs restore natural sensation to individuals with lower-limb amputations. J. Neural Eng. 15, 056002 (2018).

33. Kim, S. et al. Behavioral assessment of sensitivity to intracortical microstimulation of primate somatosensory cortex. Proc. Natl. Acad. Sci. U. S. A. 112, 15202–15207 (2015).

34. Petersen, B., Sparto, P. J. & Fisher, L. E. Clinical measures of balance and gait cannot differentiate somatosensory impairments in people with lower-limb amputation. medRxiv (2022).

35. Jones, M. G. et al. Neuromodulation using ultra low frequency current waveform reversibly blocks axonal conduction and chronic pain. Sci. Transl. Med. 13, eabg9890 (2021).

36. Saal, H. P. & Bensmaia, S. J. Biomimetic approaches to bionic touch through a peripheral nerve interface. Neuropsychologia 79, 344–353 (2015).

37. Okorokova, E. V, He, Q. & Bensmaia, S. J. Biomimetic encoding model for restoring touch in bionic hands through a nerve interface. J. Neural Eng. 15, 66033 (2018).

38. Valle, G. et al. Biomimetic Intraneural Sensory Feedback Enhances Sensation Naturalness, Tactile Sensitivity, and Manual Dexterity in a Bidirectional Prosthesis. Neuron 100, 37-45.e7 (2018).

39. A., G. J. et al. Biomimetic sensory feedback through peripheral nerve stimulation improves dexterous use of a bionic hand. Sci. Robot. 4, eaax2352 (2019).

40. Nanivadekar, A., Chandrasekaran, S., Gaunt, R. & Fisher, L. RNEL PerceptMapper. (2020) doi:10.5281/ZENODO.3939658.

41. Prieto, T. E., Myklebust, J. B., Hoffmann, R. G., Lovett, E. G. & Myklebust, B. M. Measures of postural steadiness: differences between healthy young and elderly adults. IEEE Trans. Biomed. Eng. 43, 956–966 (1996).

42. Cripps, A., Livingston, S. C. & DeSantis, B. M. The Test-Retest Reliability and Minimal Detectable Change of the Sensory Organization Test and Head-Shake Sensory Organization Test. in (2016).

43. Wrisley, D. M., Marchetti, G. F., Kuharsky, D. K. & Whitney, S. L. Reliability, Internal Consistency, and Validity of Data Obtained With the Functional Gait Assessment. Phys. Ther. 84, 906–918 (2004).

44. Beninato, M., Fernandes, A. & Plummer, L. S. Minimal Clinically Important Difference of the Functional Gait Assessment in Older Adults. Phys. Ther. 94, 1594–1603 (2014).

45. Pesarin, F. & Salmaso, L. The permutation testing approach: a review. Statistica 70, 481–509 (2010).

